# Predictive value of Altmetric Score on prospective citation and bibliometric impact: rise of a new argot

**DOI:** 10.1101/2020.05.15.20102830

**Authors:** David BT Robinson, Arfon GMT Powell, Jennifer Waterman, Luke Hopkins, Osian P James, Richard J Egan, Wyn G Lewis

**Affiliations:** Health Education and Improvement Wales’ School of Surgery, Ty Dysgu, Cefn Coed, Nantgarw, CF15 7QQ; Division of Cancer & Genetics, Cardiff University, Heath Park, Cardiff, CF14 4XW; Morriston Hospital, Cwmrhydceirw, Swansea, SA6 6NL; Swansea University, Singleton Park, Sketty, Swansea, SA2 8PP

**Keywords:** Bibliometrics, Citation, Altmetrics, Surgery

## Abstract

**Background:** Bibliometric and Altmetric analyses provide important but alternative perspectives regarding research article impact. This study aimed to establish whether Altmetric Score (AS) was associated with citation rate, independent of bibliometrics.

**Method:** Citations for a previously reported cohort of 100 most cited articles associated with the keyword “Surgery” (2018, Powell *et al*), were collected and a three-year interval Citation Gain (iCG) evaluated. Previous citation count, Citation Rate Index (CRI), AS, five-year Impact Factor, and Oxford Centre for Evidence Based Medicine (OCEBM) levels were used to classify citation rate prospect.

**Results:** During follow-up, the median iCG was 161 (IQR 83–281), with 73 and 62 articles receiving an increase in CRI and AS, respectively. Median CRI and AS increase were 2.8 (−0.1–7.7) and 3 (0–4), respectively. Receiver-Operator-Characteristic (ROC) analysis revealed that CRI (AUC 0.86 (95% CI 0.79–0.93), p<0.001) and AS (Area Under Curve (AUC) 0.65 (95% CI 0.55–0.76), p=0.008) were associated with higher iCG. AS critical threshold ≥ 2.0 was associated with better iCG when dichotomised at iCG median (OR=4.94, 95% CI 1.99–12.26, p=0.001) and iCG Upper Quartile (UQ, OR=4.13, 95% CI 1.60–10.66, p=0.003). Multivariable analysis identified that only CRI was independently associated with iCG when dichotomised at the median (OR 18.22, 95% CI 6.70–49.55, p<0.001) and UQ (OR 19.30, 95% CI 4.23–88.15, p<0.001).

**Conclusion:** Citation Rate Indices and Altmetric Scores are important predictors of interval Citation Gain, and better at predicting future citations than the historical and established Impact Factor and OCEBM quality of evidence descriptors.

## Introduction

Impact and academic reach, are sought after by authors and journal editorial boards alike. Presently, the number of citations achieved by any given article is the most commonly used measure of academic reach, and is the foundation of author Hirsch Index (h-index) and journal Impact Factor (IF). Moreover, the UK’s Research Excellence Framework (REF), which evaluates the quality of the research performed by higher education institutions, allocates funding according to academic outputs associated with journals held in high esteem on the basis of these traditional measures_1,2_. But the scientific publishing climate is changing, with a fresh argot rising, allied to novel and more immediately accessible metrics. Consequently, the concept of what constitutes impact remains as important and relevant a philosophical question as ever_3,4_.

Citations take time to accumulate, and other faster assessment means have arisen resulting in alternative metrics, or ‘Altmetrics’. These extend the concept of citation beyond mention in other scholarly articles; by recording for example, how often an article is downloaded, and applies to people, journals, books, data sets, presentations, videos, source code repositories, and web pages. The Altmetric Score (AS) calculates impact based on diverse on-line research outputs, including social media, blogs, on-line news media, and on-line reference managers_3_.

Articles with higher AS have been reported to be associated with higher citation counts3,5, but whether this symbolises preliminary intensified curiosity, or sustained citation growth is unknown. The aim of this study was to identify the factors associated with future citation accumulation, using a historical control cohort of the 100 most cited articles in surgery (2017)3. The hypothesis was that AS is directly proportional to prospective citations and consequently represents an important indicator of citation trajectory.

## Methods

Citation counts related to a previously reported cohort of 100 all-time most cited articles ring-fenced from a search performed by *Powell et al in 2017*, and associated with the keyword “surgery” were re-analysed3. Thomas Reuters’ Web of Science citation index was again accessed, providing a three-year interval Citation Gain (iCG). Previously collected bibliometric data, AS, citation count, Citation Rate Index (CRI – the total number of citations divided by the number of years since the article was published)6, five-year Impact Factor (IF), and Oxford Centre for Evidence Based Medicine (OCEBM)7 quality of evidence levels were used to classify citation rate prediction.

### Statistical analysis

All data were expressed as median (interquartile range (IQR)) and non-parametric inferential statistical methods used throughout. The OCEBM scoring system for quality of evidence, were reported in a Likert scale format; Spearman correlation coefficient test was used to test the relationship between quality of evidence and iCG. The iCG was clustered into deciles to meet the test assumptions. Receiver-Operator-Characteristic (ROC) analysis assessed the predictive value of continuous variables with the primary outcome measure of iCG dichotomized at the median and top quartile8. Univariable (UVA) and multivariable (MVA) logistic regression models were developed to identify independent associations with the primary outcome measures. Variables with a p-value of < 0.100 following UVA were included in the MVA, using a backward conditional model. All statistical analysis was performed using SPSS Statistics, Version 25.0 (IBM Corp. Armonk, NY: IBM Corp.).

## Results

The total number of citations and iCG for the cohort was 100,127 and 28,990 respectively. Every article received more citations with a median iCG of 161 (IQR 83–281). Seventy-three articles had an increase in CRI (median gain 2.8 (IQR-0.1–7.7)) and 62 articles an increase in AS (median gain 3 (IQR 0–4)). The top 10 articles related to the highest iCG can be found in table 1_9–18_, and the full article list is available in supplementary table 1. Of the original 10 most cited articles in 2017, eight maintained their top ten status three years later related to total citations.

**Table 1:**
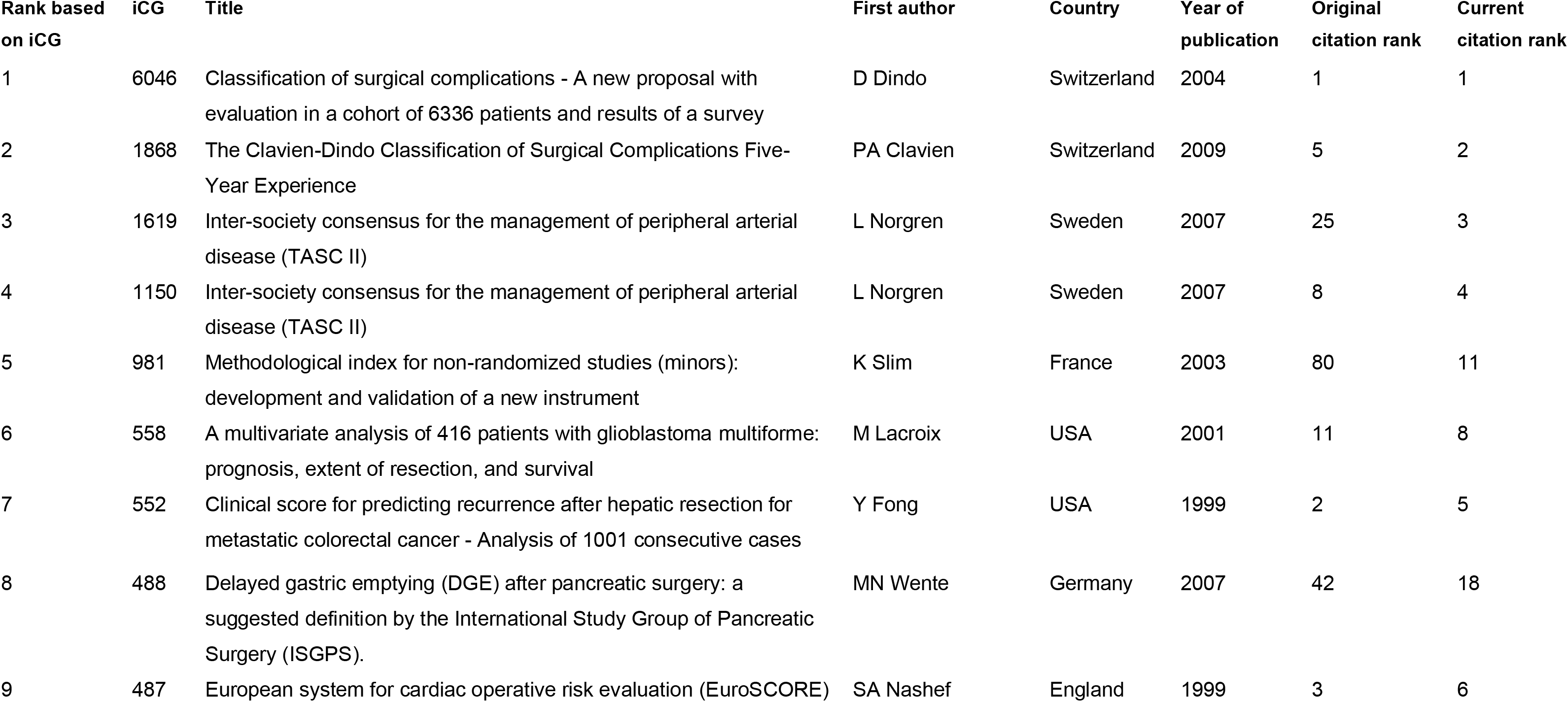
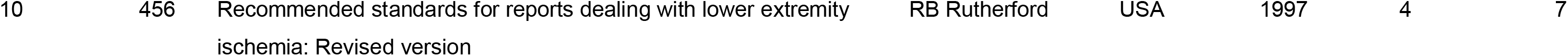
Top ten articles based on iCG

### Bibliometric factors associated with iCG

Median article iCG when first or last authors featured more than once, was 240 (IQR 128–701.5), compared with 153.5 (IQR 75.5–257, p=0.011) when authors featured only once in the 100 most cited articles. Multicentre studies were not associated with higher iCGs compared with single centre studies (median 183 (IQR 98–336) vs. 143 (IQR 80–257), p=0.209). Median iCG related to the country of the leading institute, OCEBM levels, research subject, disease category, and specialty can be found in supplementary figures 1–5. The highest and lowest iCG related to country, OCEBM level, subject, disease category and specialty were Switzerland vs. Japan (median iCG 1084 (IQR 251–5002) vs. 57 (IQR 42–134)), level 5 vs. level 3 (291 (IQR 98–456) vs. 110 (IQR 83–135)), epidemiology vs. nutrition (456 (IQR 291–719) vs. 101 (IQR 101–101)), musculoskeletal vs. respiratory (222 (IQR 166–321) vs. 54 (IQR 54–54)) and vascular vs. plastic surgery (235 (74–977) vs. 48 (IQR 48–48)), respectively.

The relationship between the original AS, number of citations, CRI, and five-year journal IF related to iCG, can be found in figure 1. Linear regression analysis identified that original citation count (r_2_=0.797, 95% CI 0.90–1.10, p<0.001) and CRI (r_2_=0.908, 95% CI 12.47–14.17, p<0.001) were associated with iCG. AS (r_2_=0.024, 95% CI –3.40–28.66, p=0.121), five-year journal IF (r_2_= 0.011, 95% CI –25.82–80.24, p=0.311), and OCEBM level (r_2_ = 0.002, 95% CI –124.50–78.45, p=0.653), were not associated with iCG. When two outliers were removed from the AS analysis (figure 1b), AS (r_2_ = 0.199, 95% CI 0.01–0.01, p<0.001) was also associated with iCG.

**Figure 1:**
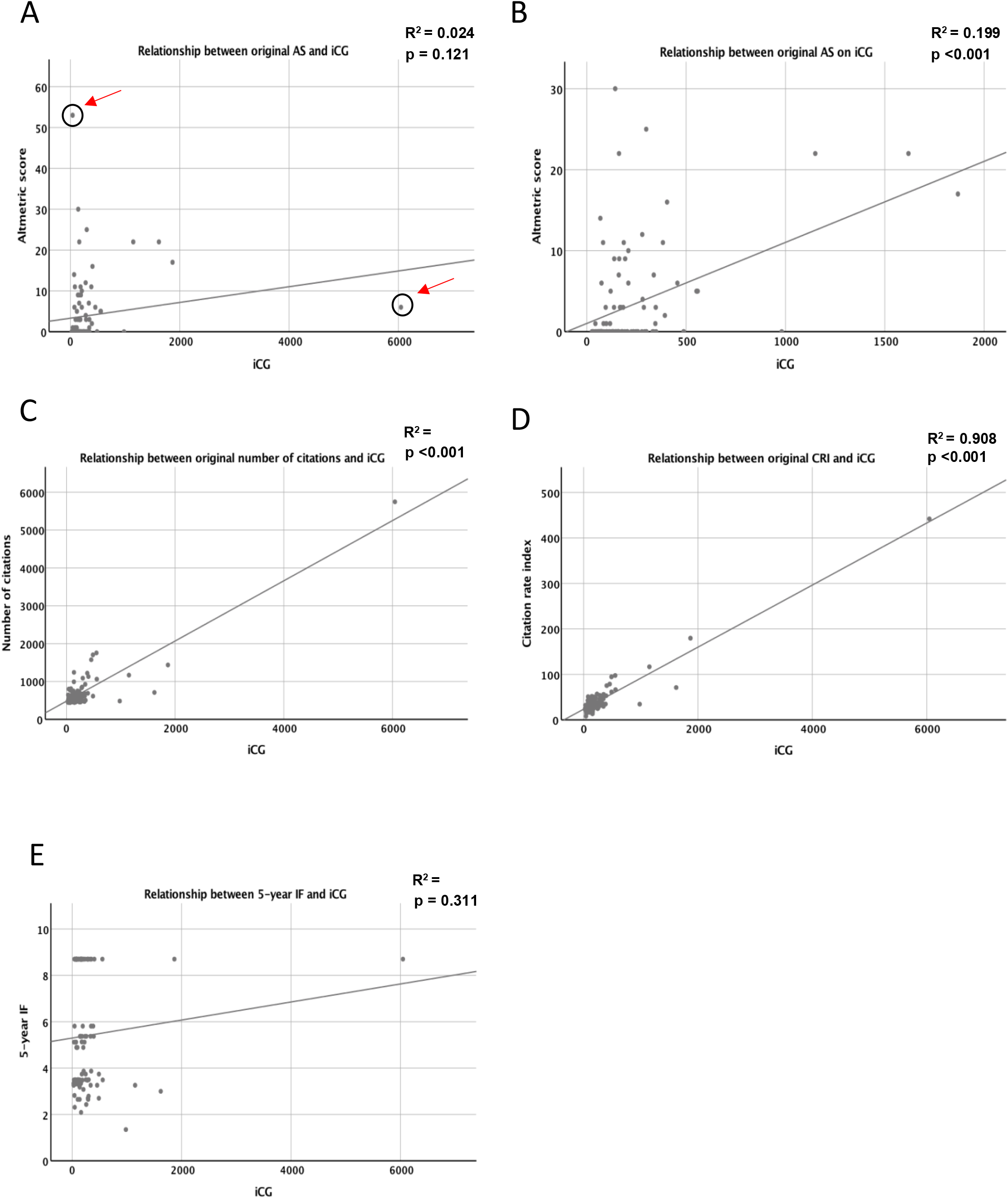
Scatter plots illustrating the relationship between the original variables and the increase in the number of citations over time. **A**:The relationship between original AS and iCG. **B**: The relationship between original AS and iCG with the two outliers removed – as marked in A. **C**: The relationship between original citation count and iCG. **D**: The relationship between original CRI and iCG. **E**: The relationship between original 5-year IF and iCG.

### Creation of dichotomisation thresholds for confounding adjustment

Receiver-Operator-Characteristic (ROC) curve analysis was used to identify possible critical thresholds for variables associated with higher iCG (Figure 2). Table 1 outlines the critical thresholds that were associated with higher iCG. No definitive critical thresholds were identified for five-year IF (figure 2d) or OCEBM levels (figure 2e). The critical thresholds for original AS, citation count, and CRI were 1.5, 520.5 and 34.4, respectively. Articles with AS > 2 (median iCG 133 (IQR 71–234.5) vs. 209 (IQR 143–393), p<0.001), citation count > 521 (120 (IQR 70–173) vs.192 (IQR 116–300), p=0.004) and CRI > 34.4 (95 (IQR 61–157) vs. 250 (IQR 163–383), p<0.001) were associated with higher median iCGs.

**Figure 2:**
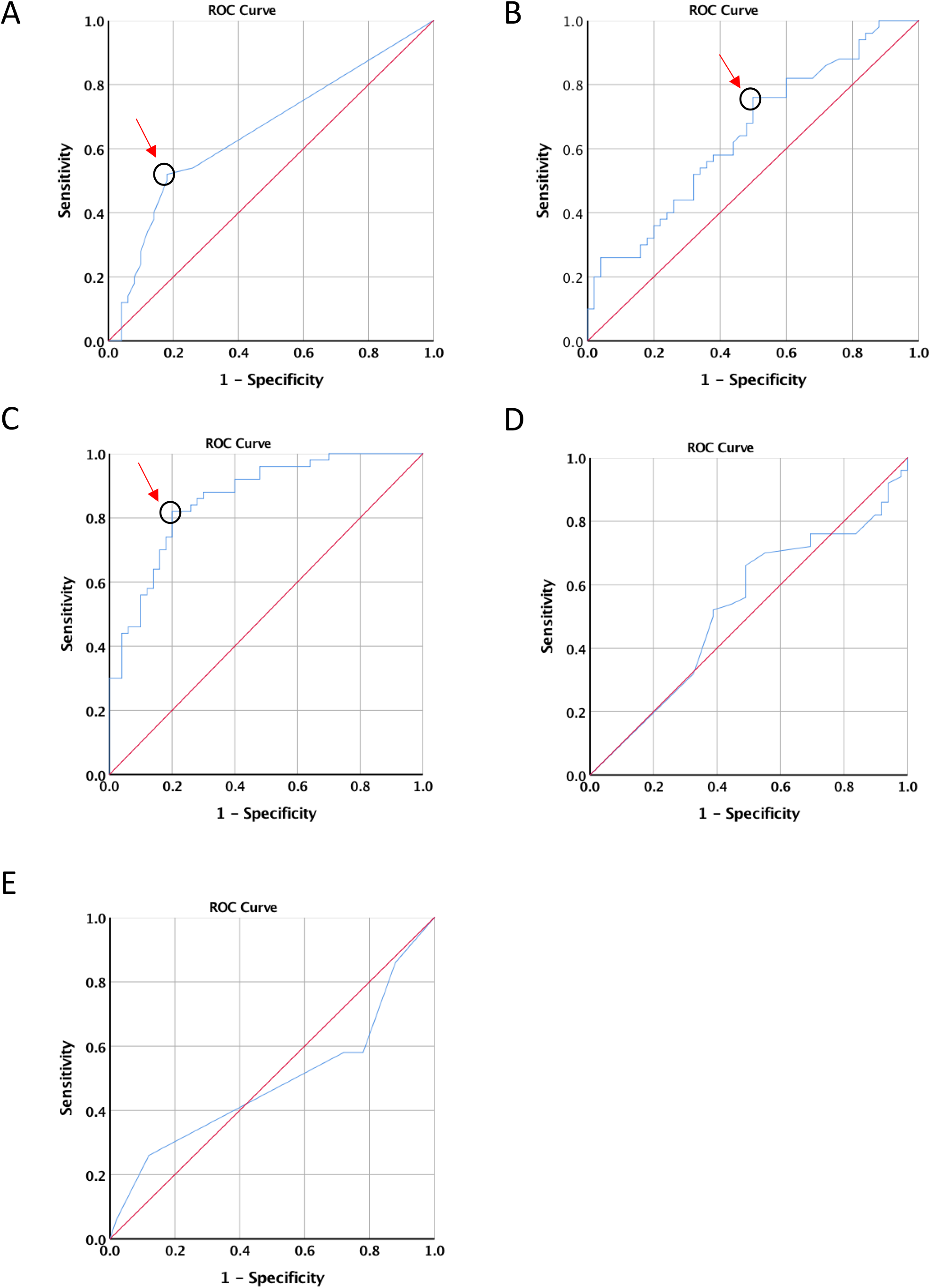
ROC curves to identify critical thresholds for variables. ROC curves associated with A: Original AS. B: Original citation count. C: Original CRI. D: Original 5-year IF. E: OCEBM level. Arrows and circles indicate the point taken to establish critical thresholds

**Table 2:** The relationship between bibliometric variables, CRI and AS in predicting gains in citations.

|  | **Median score** | **IQ range** | **AUC** | **95% CI** | **p-value** | **Threshold** |
| --- | --- | --- | --- | --- | --- | --- |
| **Original AS** | 0 (0-53) | 0-5 | 0.65 | 0.55-0.76 | 0.008 | 1.5 |
| **Original citation** | 573.5 (446-5746) | 488.8-707.5 | 0.65 | 0.54-0.75 | 0.012 | 520.5 |
| **Original CRI** | 34.7 (8.0-442) | 26.3-45.9 | 0.86 | 0.79-0.93 | <0.001 | 34.4 |
| **Original 5-year IF** | 4.9 (1.35-8.70) | 3.3-8.7 | 0.52 | 0.41-0.64 | 0.680 | N/A* |
| **OCEBM** | N/A | N/A | 0.46 | 0.35-0.58 | 0.545 | N/A* |

\* The data assumptions of the Youden index were violated, therefore dichotimisation points associated with original IF and quality of evidence were not considered.

### Univariable and Multivariable analysis of factors associated with above the median and upper quartile iCG

Univariable and multivariable analyses of factors associated with iCG can be found in table 3. On univariable analysis, multicentre studies (p=0.023), articles where the first or last author featured more than once in the 100 most cited articles (p=0.044), original AS > 2 (p=0.001), original citation count > 521 (p=0.008), and original CRI > 34.4 (p<0.001) were associated with higher iCG dichotomised around the median. When dichotomised around the upper quartile, articles where the first or last author featured more than once (p=0.041), original AS > 2 (p=0.003), original citation count > 521 (p=0.048), and original CRI > 34.4 (p<0.001), were associated with higher iCG. On multivariable analysis only CRI was associated with higher citation accrual when iCG was dichotomised around the median (OR 18.22 (95% CI 6.70–49.55), p<0.001), and upper quartile (OR 19.30 (95% CI 4.23–88.15), p<0.001).

**Table 3:**
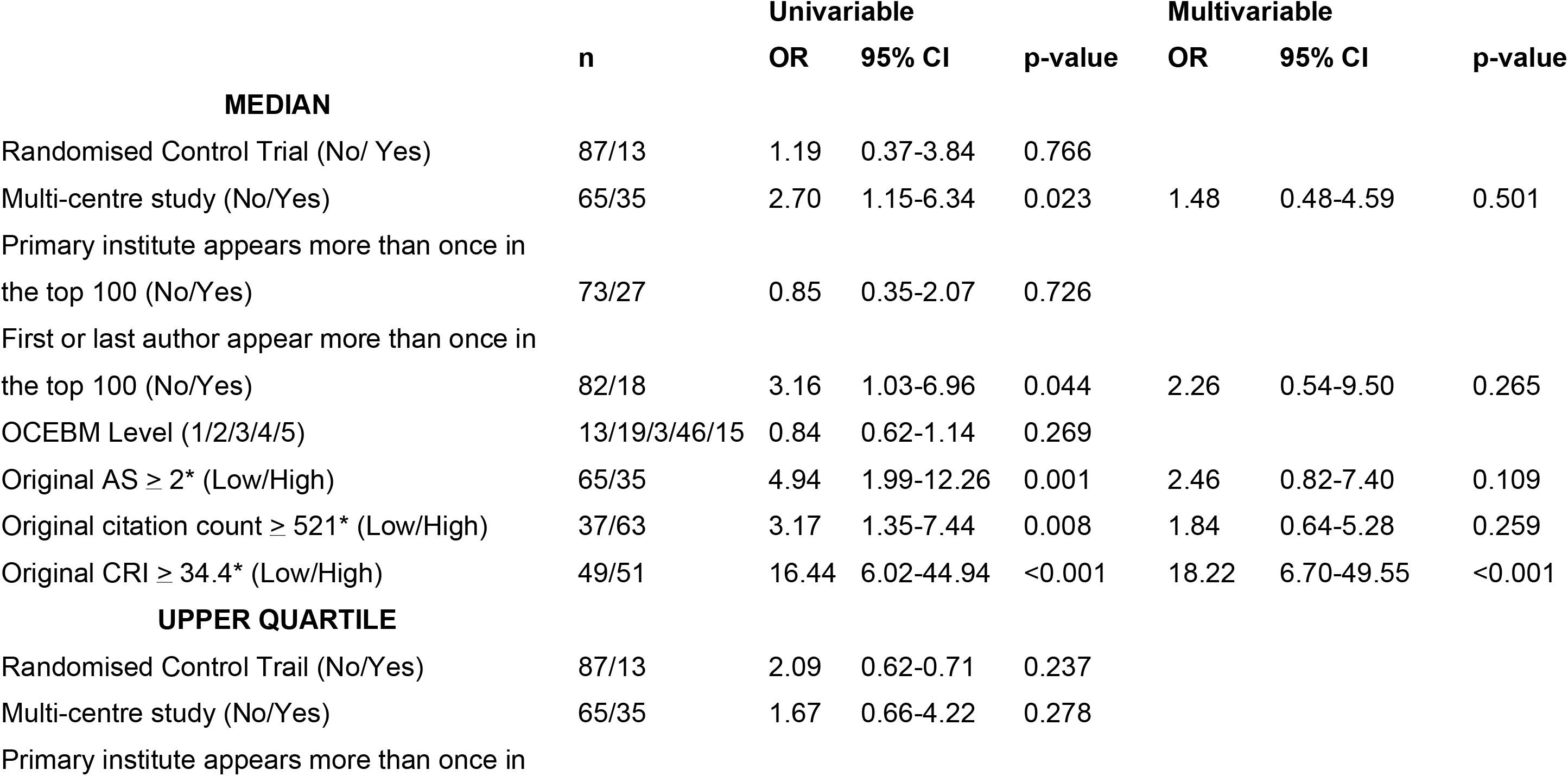
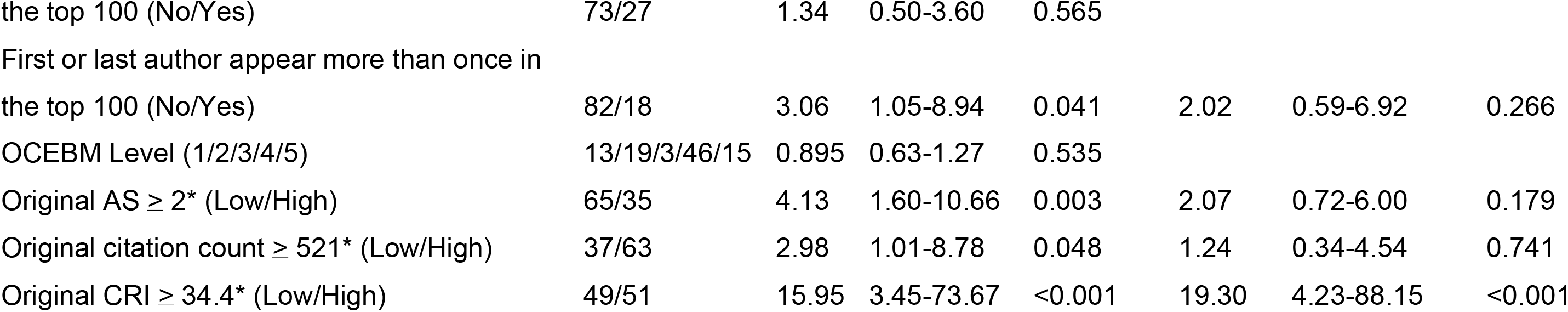
Univariable and multivariable analysis of factors associated with higher iCG. Results pertain to binary logistic regression tests. OR = Odds Ratio; CI = Confidence Interval; AS = Altmetric Score; CRI = Citation Rate Index. ^*^Identified from ROC curve analysis.

## Discussion

The principal finding of this study was that articles that amass citations at faster rates, continue to grow their citation counts, independent of other bibliometric parameters such as study type, evidence quality, author affiliation, and author success. Articles with a CRI greater than 34.4 were approximately 18 times more likely to accrue a greater number of citations. Altmetric Score was also associated with a higher iCG, with AS of two or more conferring a five-fold multiplication factor with respect to citation accrual over three years, however this was not independent of CRI, supporting the null hypothesis. OCEBM levels and other bibliometric parameters were not independently associated with iCG and more work to examine the factors associated with a lift in article citations is needed.

Despite common use of several impact metrics to gauge the professional success of academics, just how scientific power rises and evolves remains indistinct. Sinatra *et al*, from Boston, quantified the changes in impact and productivity of an academic throughout a career in science, and reported that impact, as measured by influential publications, was distributed randomly through a scientist’s publication sequence._19_ This random-impact rule allows the formulation of a stochastic model that uncouples the effects of individual ability, productivity, and luck, and implies the existence of universal patterns governing the rise of scientific success. The model assigns a unique individual and stable parameter Q to each scientist, which accurately predicts the evolution of a scientist’s impact, from the h-index to cumulative citations, and independent acknowledgments such as prizes. This suggests that a scientist can influence an articles impact, independent of the content of the article. This is supported by the observation in this study that authors who have more than one entry in the 100 most cited articles go on to accrue more interval citations, independent of the author’s academic affiliations_9,10,11,12,15,15,20–32_. Moreover, possessing multiple publications in this cohort was not independent of CRI in predicting iCG on statistical modelling, supporting a close association between author influence and citation accrual. Certain authors, or teams of authors, appear to be associated with a positive multiplication coefficient or constant, which when allied to a favourable project that has novelty, good fit, and timeliness, can accelerate and boost citation trajectory markedly.

Wang *et al*, from Boston, reported a mathematically derived mechanistic model, that showed that ultimate impact, characterised by the lifetime number of citations received by a manuscript, is largely predicted by a single factor, fitness_33_. Fitness is the perceived novelty and relevance that an article enjoys from the research community at large. This may explain why the Clavien-Dindo Classification article_9_, which received the greatest number of citations and had the highest iCG in this study, was one of the top ranking article in a 100 most-cited analysis of surgery_34_, visceral surgery_35_, and laparoscopic surgery_36_. When Wang *et al* compared articles from journals with an impact factor of 3.26, 10.48, and 33.62, they reported that articles with similar fitness values obtained their initial citation load in a journal dependent manner, but ultimately received similar lifetime citation counts. This may explain why journal IF was not a predictor of citation count in the original 2017 study_3_, or iCG in this study. Furthermore, the findings described by Wang *et al*, also explains how the immediacy of citation gain, which increases the IF of a journal, is supported by social media platforms. But whether the ‘social media’ impact simply pushes the mean citation load to the left, or boots overall citation counts and impact is opaque_33_.

With increasing competition for research funding and tenure, emphasis has now moved towards demonstrating impact using metrics. Historically, article citation counts have always been synonymous with impact. Researcher level *impact metrics*, which are also based on citation counts, include the Hirsch-Index, r-index, m-index, g-index, and i10 index among others._4_ Metric use is further supported by the present study’s findings, and in particular that CRI, derived from citation count, was associated independently with iCG. Academic reach is now facilitated by smart phone and electronic device use, and journals have embraced this paradigm shift with a greater online presence. Zerrweck *et al*, from Mexico City, reported that 31.5% (51.8% recreational use) of general surgeons, and 39.7% (68.6% recreational use) of bariatric surgeons use social media on a daily basis for academic purposes._37_ It is therefore possible for researchers to play an active role in the dissemination and reach of their work, which will likely result in more citations. The introduction of alternative metrics heralds the beginning of a new way of measuring impact. What remains to be seen is whether disseminating pre-print articles on social media, or in pre-print repositories, results in higher AS and citation counts; providing journal editors with early impact metrics.

This study has a number of potential inherent limitations. Articles included were published at different time points and were therefore at different stages of their citation accrual. Furthermore, original AS were not recorded at identical follow-up time points. Nevertheless, AS and CRI were associated with a higher iCG and therefore irrespective of the recorded time, these bibliometric markers are strongly associated with ongoing citation accrual and impact. Citations carry inherent bias in the form of institutional or self-citation that cannot be identified from a database search. In contrast, the study has a number of strengths. Article citations and AS were recorded simultaneously and from the premier reputable database; Thomas Reuteurs Web of Science. Moreover, the bibliometric analysis has statistical power, limiting confounding variables, and time from publication bias.

Despite the perceived reciprocal attraction between high-quality research articles and higher impact factor journals, it appears that article fitness, social media promotion, and author authority, are all important predictors of prospective article impact and citation accrual. If journal editorial strategy is to publish articles expected to acquire high numbers of citations, it may be contended that early upload to one of the emerging publishing repository web-sites, prior to formal journal submission should be encouraged. Such tactics would allow a period of attention analysis, in the form of AS and early citation accrual, prior to periodical publication.

## Data Availability

If raw data is required, requests can be made to the corresponding author.

## Declarations of interest

None. Not previously presented to a society or meeting

## Ethical Approval

Not applicable

## Funding

OP James was supported by a Joint Surgical Research Fellowship from the Royal College of Surgeons England and Health Education and Improvement Wales

## Short Title

Do Altmetrics predict future citation potential?

## Notes

### Competing Interest Statement

The authors have declared no competing interest.

## References

1 Greenhalgh T, Raftery J, Hanney S, Glover M. Research impact: A narrative review. BMC Med. 2016; 14: Article 78.

2 Building on Success and Learning from Experience. An Independent Review of the Research Excellence Framework. [Internet]. 2016. Available from: https://assets.publishing.service.gov.uk/government/uploads/system/uploads/attachment_data/file/541338/ind-16-9-ref-stern-review.pdf

3 Powell, AGMT, Bevan V, Brown C, Lewis WG. Altmetric Versus Bibliometric Perspective Regarding Publication Impact and Force. World J Surg. 2018; 42: 2745–2756.

4 Robinson, DBT, Hopkins L, Brown C, Abdelrahman T, Powell AG, Egan RJ, et al. Relative Value of Adapted Novel Bibliometrics in Evaluating Surgical Academic Impact and Reach. World J Surg. 2019; 43: 967–972.

5 Elmore SA. The Altmetric Attention Score: What Does It Mean and Why Should I Care? Toxicol Pathol. 2018; 46: 252–255.

6 Powell, AGMT, Hughes DL, Wheat JR, Lewis WG. The 100 most influential manuscripts in gastric cancer: A bibliometric analysis. Int J Surg. 2016; 28: 83–90.

7 OCEBM Levels of Evidence Working Group, Durieux N, Pasleau F, Howick J. The Oxford 2011 Levels of Evidence. Group [Internet]. 2011; Available from: https://www.cebm.net/2016/05/ocebm-levels-of-evidence/

8 Youden WJ. Index for rating diagnostic tests. Cancer. 1950 Jan; 3: 32–35.

9 Dindo D, Demartines N, Clavien PA. Classification of surgical complications: A new proposal with evaluation in a cohort of 6336 patients and results of a survey. Ann Surg. 2004; 240: 205–213.

10 Clavien PA, Barkun J, De Oliveira ML, Vauthey JN, Dindo D, Schulick RD, et al. The clavien-dindo classification of surgical complications: Five-year experience. Ann Surg. 2009; 250: 187–196.

11 Norgren L, Hiatt WR, Bell K, Nehler MR, Harris KA, Fowkes FGR, et al. Inter-Society Consensus for the Management of Peripheral Arterial Disease (TASC II). Eur J Vasc Endovasc Surg. 2007; 33: S1–S75.

12 Norgren L, Hiatt WR, Dormandy JA, Nehler MR, Harris KA, Fowkes FGR. Inter-Society Consensus for the Management of Peripheral Arterial Disease (TASC II). J Vasc Surg. 2007; 45: S5–S67.

13 Slim K, Nini E, Forestier D, Kwiatkowski F, Panis Y, Chipponi J. Methodological index for non-randomized studies (Minors): Development and validation of a new instrument. ANZ J Surg. 2003; 73: 712–716.

14 Lacroix M, Abi-Said, D, Fourney DR, Gokaslan ZL, Shi W, DeMonte, F, et al. A multivariate analysis of 416 patients with glioblastoma multiforme: Prognosis, extent of resection, and survival. J Neurosurg.2001 95: 190–198.

15 Fong Y, Fortner J, Sun RL, Brennan MF, Blumgart LH. Clinical score for predicting recurrence after hepatic resection for metastatic colorectal cancer: Analysis of 1001 consecutive cases. Ann Surg. 1999; 230: 309–318.

16 Wente MN, Bassi C, Dervenis C, Fingerhut A, Gouma DJ, Izbicki JR, et al. Delayed gastric emptying (DGE) after pancreatic surgery: A suggested definition by the International Study Group of Pancreatic Surgery (ISGPS). Surgery. 2007; 142: 761–768.

17 Nashef, SAM, Roques F, Michel P, Gauducheau E, Lemeshow S, Salamon R. European system for cardiac operative risk evaluation (EuroSCORE). Eur J Cardio-thoracic Surg. 1999; 16: 9–13.

18 Rutherford RB, Baker JD, Ernst C, Johnston KW, Porter JM, Ahn S, et al. Recommended standards for reports dealing with lower extremity ischemia: Revised version. J Vasc Surg. 1997; 26: 517–538.

19 Sinatra R, Wang D, Deville P, Song C, Barabási, A-L. Quantifying the evolution of individual scientific impact. Science (80-). 2016; 354.

20 Fong Y, Sun RL, Jarnagin W, Blumgart LH. An analysis of 412 cases of hepatocellular carcinoma at a Western center. Ann Surg. 1999; 229: 790–799.

21 Strasberg SM, Hertl M, Soper NJ. An analysis of the problem of biliary injury during laparoscopic cholecystectomy. J Am Coll Surg. 1995; 180: 101–125.

22 Heald RJ, Husband EM, Ryall, RDH. The mesorectum in rectal cancer surgery—the clue to pelvic recurrence? Br J Surg. 1982; 69: 613–616.

23 Heald RJ, Moran BJ, Ryall, RDH, Sexton R, yall, RDH, Sexton, R MacFarlane JK. Rectal Cancer: The Basingstoke experience of total mesorectal excision, 1978–1997. Arch Surg.1998 133: 894–899.

24 Adam R, Delvart V, Pascal G, Valeanu A, Castaing D, Azoulay D, et al. Rescue surgery for unresectable colorectal liver metastases downstaged by chemotherapy: A model to predict long-term survival. Ann Surg. 2004; 240: 644–657.

25 Schauer PR, Ikramuddin S, Gourash W, Ramanathan R, Luketich J. Outcomes after laparoscopic Roux-en-Y gastric bypass for morbid obesity. Ann Surg. 2000; 232: 515–529.

26 Schauer PR, Burguera B, Ikramuddin S, Cottam D, Gourash W, Hamad G, et al. Effect of Laparoscopic Roux-En Y Gastric Bypass on Type 2 Diabetes Mellitus. Ann Surg. 2003; 238: 467–485.

27 Clavien PA, Sanabria JR, Strasberg SM. Proposed classification of complications of surgery with examples of utility in cholecystectomy. Surgery. 1992; 111: 518–526.

28 Kassell NF, Torner JC, Jane JA, Haley EC, Adams HP. The International Cooperative Study on the Timing of Aneurysm Surgery. Part 2: Surgical results. J Neurosurg. 1990; 73: 37–47.

29 Kassell NF, Torner JC, Haley EC, Jane JA, Adams HP, Kongable GL. The International Cooperative Study on the Timing of Aneurysm Surgery. Part 1: Overall management results. J Neurosurg. 1990; 73: 18–36.

30 Kehlet H, Wilmore DW. Evidence-based surgical care and the evolution of fast-track surgery. Ann Surg. 2008; 248: 189–198.

31 Kehlet H, Wilmore DW. Multimodal strategies to improve surgical outcome. Am J Surg. 2002; 183: 630–641.

32 Bismuth H, Adam R, Lévi F, Farabos C, Waechter F, Castaing D, et al. Resection of nonresectable liver metastases from colorectal cancer after neoadjuvant chemotherapy. Ann Surg. 1996; 224: 509–520.

33 Wang D, Song C, Barabási, AL. Quantifying long-term scientific impact. Science (80-). 2013; 342: 127–132.

34 Manuel Vázquez, A, Latorre Fragua R, López Marcano A, ópez Marcano A Ramiro Pérez, C, Arteaga Peralta V, de la Plaza-Llamas, R, et al. The Top 100: A Review of the Most Cited Articles in Surgery. Cirugía Española.2019 97: 150–155.

35 Müller, M, Gloor B, Candinas D, Malinka T. The 100 Most-Cited Articles in Visceral Surgery: A Systematic Review. Dig Surg. 2016; 33: 509–519.

36 Mellor KL, Powell, AGMT, Lewis WG. Laparoscopic Surgery’s 100 Most Influential Manuscripts: A Bibliometric Analysis. Surg Laparosc Endosc Percutaneous Tech. 2018; 28: 13–19.

37 Zerrweck C, Arana S, Calleja C, Rodríguez N, Moreno E, Pantoja JP, et al. Social media, advertising, and internet use among general and bariatric surgeons. Surg Endosc. 2020; 34: 1634–1640.

